# Seizure Forecasting with Multiple Time Scales and Features

**DOI:** 10.1101/2025.06.02.25328687

**Authors:** Yueyang Liu, Artemio Soto-Breceda, Philippa Karoly, David B. Grayden, Mark J. Cook, Dean R. Freestone, Daniel Schmidt, Levin Kuhlmann

**Affiliations:** Dept. of Data Science and AI, Faculty of Information Technology, Monash University; Dept. of Biomedical Engineering and Graeme Clark Institute, The University of Melbourne; Dept. of Medicine, St Vincent’s Hospital, The University of Melbourne; Seer Medical Pty, Melbourne, Australia

**Keywords:** epileptic seizure forecasting, dynamical systems, critical slowing down, neural filtering, EEG feature analysis

## Abstract

Forecasting epileptic seizures is a difficult task. Studies of seizure prediction have investigated many different EEG features, but none of them have been useful enough to be applied in clinical practice beyond clinical trials. Moreover, most of these features have been applied to short-term intracranial EEG (iEEG) recordings, limiting the possibility of reliable statistical evaluation. Recent work has shown that seizure forecasting using multiple long time scale cycle analysis (from 40 min to 1 month) and with long-term recordings (>6 months duration) provides the most statistically robust methods.

This paper investigates a large subset of features from the past and present to unravel which features and feature analysis methods will yield the best performance on long-term iEEG recordings (from 14 patients with focal epilepsy) and thus provide the most reliable step toward clinical utility. Specifically, this study implements a multiple long-time scale cycle feature analysis framework for seizure forecasting that considers the state-of-the-art time series features of critical slowing down (autocorrelation and variance) as well as interictal epileptiform discharge (IED) / spike rate, High Frequency Activity (HFA), seven different univariate features, and three Neural Mass Model (NMM) features based on brain dynamics.

Seizure phase histograms of all the features are then analyzed to investigate each feature’s potential for seizure forecasting by evaluating corresponding synchronization indices (SI) on fast (40 minutes to 2 days) and slow (2+ days) wideband time scales. This analysis shows that, for different patients, different subsets of features had SI values exceeding 0.7, indicating their utility for seizure forecasting and suggesting a multi-feature selection approach is required. Out of all combinations considered, the overall performance comparison across patients highlights that ‘autocorrelation + variance + NMM + spike rate’ features achieve the highest average AUC of 0.83, showcasing its performance in forecasting seizures.

In conclusion, a model is proposed that has a similar performance compared to the state-of-the-art method, without the need of selecting the best channel prior to model building. Light is also shed on the comparative performance on long-term recordings of many of the seizure forecasting features considered in the past.

## Introduction

Epilepsy is a common neurological disorder characterized by the occurrence of recurrent seizures, which is estimated to affect about one percent of the world population ^1^. Among these patients, about one third of them cannot be treated with either medication or surgery ^2^. As an alternative, seizure forecasting could offer significant benefits by enabling individuals to seek assistance or avoid hazardous situations.

The mechanism of the transition between normal state to a seizure state is not fully understood ^3^. Understanding this mechanism could be helpful in generating new forecasting and treatment methods and is an important direction of research that parallels more generic signal feature-based approaches.

Many researchers have investigated a variety of features for seizure prediction or forecasting ^2,3^. While a variety of comparison studies of different features have been done on short-term iEEG recordings (< 2 weeks duration) ^4–8^ as they are easier to obtain, these studies lack the necessary amount of data per patient to robustly statistically assess performance of these methods. This is because seizures are rare events and therefore large amounts of data are needed to perform robust statistical analysis. Various studies using long-term iEEG recordings have demonstrated robust seizure forecasting performance using different features or approaches ^9– 16^, but no study as yet has completed a more comprehensive evaluation of the seizure forecasting performance of different features and their combinations on long-term recordings. This paper seeks to fill this gap.

Of the methods considered on long-term recordings that also seek to provide a mechanistic understanding, an approach based on critical slowing down has achieved state of the art performance ^13^. The analysis framework associated with this approach is employed in this paper both as a benchmark and as a scaffold for performing the methods comparison. The framework relies on the analysis of iEEG features analyzed on fast (40+ minutes to 2 days) and slow (2+ days) wideband time scales.

Critical slowing down is a phenomenon of a dynamical system approaching a critical transition. The phenomenon refers to the tendency of a dynamical system to take longer to return to the equilibrium point after perturbations ^17^, and is indicated by an increase in signal variance and autocorrelation when close to a transition point. Not all abrupt transitions will be preceded by slowing down, as a transition may result from a big external impact instead ^18^. However, if a system is driven towards the transition point at a moderate pace ^19^, we can expect critical slowing to happen on certain time-scales. As such, it is a sensible measure to use for seizure forecasting.

From a mechanistic perspective, computational neural models also provide valuable insights into the dynamics of the brain and facilitate a dynamical systems viewpoint consistent with the critical slowing down approach. Researchers have suggested computational models of epilepsy, in particular neural mass models (NMMs), are able to show the change in brain state via critical transition ^20–22^. Furthermore, neurophysiological variables of these models such as population averaged synaptic connection strengths can be inferred from data to forecast seizures ^8,23^.

To complete the proposed comparison of seizure forecasting features, this paper considers these more mechanistic-interpretation oriented features of critical slowing down and NMM parameters, along with clinical features, such as IED/spike rates and HFA, and other univariate features from the past. Moreover, with the fast development of machine learning and deep learning, researchers have extended their application to the field of seizure forecasting ^12,24–27^ to yield further improvements in performance. As such machine learning is applied here to combine different subsets of features from the past and present, and evaluate and compare the seizure forecasting performances of these different combinations to yield a clearer picture of what works best and help move seizure forecasting into the realm of being clinically useful.

## Methods

### Dataset

We used the long-term iEEG dataset obtained from the first-in-man clinical trial of a seizure advisory device ^9^ (Human Research Ethics Committee, St. Vincent’s Hospital, Melbourne – approval LRR145/13). The long-term recordings (range from 6 months to 3 years) consist of 16 electrodes sampled at 400 Hz. The original study recorded 15 patients, although 14 were used in this study. Patient 9 was excluded due to their low number of seizures. Further information on the collection of the data can be found in the original trial ^9^.

### Preprocessing

To reduce the computational cost, we extracted features on a 1 second time frame for every 2 minutes, following the state-of-the-art critical slowing down-based seizure forecasting method ^13^. The first 3 s from the 2 min window were extracted and were filtered using a finite impulse response lowpass filter with a cut-off of 170 Hz to remove 200 Hz artificial noise in the data when the device was charged. After filtering, the first and third second were removed, and only the middle second was kept. Features were then calculated based on the 1 s of filtered recording for each nonoverlapping 2 min window in each recording.

### EEG Feature Extraction

A total of 14 features were calculated on the sampled data. The two critical slowing down features, Autocorrelation Function Width (ACFW) and Variance were applied ^13^. The ACFW was taken as the width at the half maximum of the autocorrelation function. Three additional features were obtained using the Jansen and Rit ^28,29^ Neural Mass Model (NMM) and a novel artificial neural network-based filter ^30^. The NMM model represents the cerebrocortical patch of neuronal tissue underlying the iEEG electrode and consists of three neural populations (see Supplementary Fig. S1) – pyramidal, excitatory stellate, and inhibitory – connected via four population averaged synaptic connections, whose strengths are labelled α_*pe*_, α_*pi*_, α_*ep*_ and α_*ip*_. The model were also driven by an external input *u*, which represents the lumped long-range inputs from other cortical areas and deep brain structures such as the thalamus. Each iEEG channel was independently fed to the filter to estimate these parameters of the NMM and the average values within a given window were used as features. Only the parameters α_*pe*_, α_*pi*_ and *u* were used for forecasting as they showed the most sensitivity to changes in data ^31,32^. Further details on the estimation of the NMM parameters are provided in the supplementary information.

Furthermore, Univariate Features were also calculated to explore how they perform on long-term recordings. Features were selected from the literature with priority given to computationally efficient features that can preserve device battery life. These include the Hjorth Parameter called Complexity ^33^, decorrelation time (decorr_time) ^4^, approximate entropy (ApEn) ^34^ and power in the delta (0.5 - 4 Hz), theta (4 – 7 Hz), alpha (7 – 12 Hz) and beta (12 - 30 Hz) bands ^35,36^. The Hjorth Parameter, Complexity, indicates the signal’s frequency content and the variation of the frequency content. Decorrelation Time is defined as the time which the first zero-crossing of the autocorrelation function occurs and is useful in characterising the temporal dynamics of the signal. Approximate Entropy measures the regularity of the signal, which indicates if more complexity or randomness is present.

High Frequency Activity (HFA) rate was also included as a feature, defined as high-frequency (80-170 Hz) events with amplitudes significantly higher than the background EEG ^37^. To improve computational efficiency and to evaluate this feature for seizure forecasting, the method for obtaining HFA has been modified from the original research ^37^. On each channel, 300 segments of 1 min data were randomly selected from the recordings of the first 50 days before filtering using a 12-order infinite impulse response (IIR) bandpass filter between 80- 170 Hz. The threshold to detect HFA was determined as the amplitude at 5 standard deviations of the Gaussian curve fitted to the histogram of the processed data ^38^. For every 2 min, a 1 min recording was filtered by the same IIR filter and passed through a Hilbert transform to extract the signal envelope. HFA was detected and counted when the signal envelope crossed the threshold for the channel ^38^. The HFA rate was the number of HFA occurrences during the minute. Be aware that, in real time, the calculation must be done after the 1 min recording, while other features are calculated after the first 3 seconds of the 1 min recording, so HFA incorporates a substantial delay.

For the final feature, a correlation-based algorithm was applied to detect Interictal Epileptiform Discharge (IEDs) or spikes. It compared the iEEG signal to a template which has been previously described and benchmarked ^32^. Around 100 candidate epileptiform spikes were detected and verified individually for every electrode in every patient by a board approved epileptologist. The template was taken as the average waveform to automatically detect epileptiform spikes. Spikes were then detected by computing the correlation between the template and a sample iEEG. Sections of iEEG with a correlation above 0.85 were considered new spikes. IED/spike rates were then obtained for each window of data.

### Multi-time scale analysis of features

The state-of-the-art seizure forecasting methods ^13,14^ are dependent on long time scales. Following this approach, “long/slow” and “short/fast” cycle time-series were obtained from each feature in a causal way, to ensure the current feature values would not be affected by future data. After the calculation of the features, a trailing moving average was applied with a window of two days to identify the “long/slow” cycle. This slow cycle was subtracted from the raw feature time series and a trailing moving average with a window of 40 minutes was applied to this result to obtain the “short/fast” cycle covering periods between 40 minutes and 2 days.

At the end, the Hilbert Transform ^39^ was implemented on both cycles to derive the signal phases. This approach provides a flexible wideband approach to detect the most optimal fast and slow cycles without having to specify which are the most optimal time scales, which is difficult to do a priori.

### Seizure tuning of feature phases on multiple time-scales

Seizure phase histograms were analysed to explore which features might be the most suitable for seizure forecasting for a given patient. Seizure phase histograms involve creating circular histograms of the Hilbert phase of a given cycle signal, where the phase from 0 to 2π is divided into *K* bins and the height of the *k*th bin is the number of seizures occurring in the phase bin. To find the features which show the highest tuning of seizures to specific phases of their fast and slow cycles, the synchronisation index (SI) was calculated^40^. The SI is given by

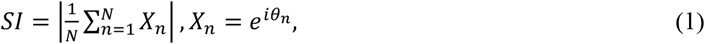

where θ_*n*_ is the phase of the cycle signal for the *n*th seizure, 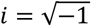, and *N* represents the total number of seizures ^13^.

An SI score close to 1 indicates the feature has higher potential to forecast seizures, as more seizures are synchronised within similar phases of the signal. On the other hand, an SI score close to 0 indicates the feature has no potential to forecast seizures, since seizures are distributed evenly across phases.

For each patient, the best channel was selected by picking the one with the highest SI score regardless of features or cycle, as we would like to keep the best feature over all time. All the other features were selected from the same channel. This analysis considered all data and did not play a role at all in the following seizure forecasting methods.

### Forecasting

The aim of forecasting is to provide warning in the 2-4 minute window before a seizure. Given potential errors in labelling seizure onset times forecasting in the 0-2 minutes before a seizure is avoided. Assume the start of a seizure is at data point *x*_*t*_; we label the data point *x*_*t*-2_ as preseizure stage and *x*_*t*-1_ and *x*_*t*_ as non-preseizure stage, where each time step is 2 min. As *x*_*t*_ is the start of the seizure which is the seizure stage, and we do not want *x*_*t*-1_ to be predicted as a warning window, we label *x*_*t*-2_ as the only preseizure stage. Training, validation, and testing were then based on these labels, meaning that only a warning provided at time *t* − 2 would be considered as a success. In a perfect scenario, the system would be silent at all times except for the preseizure stage window.

In order to initialize the training process, we chose to use the first 50 days as a start. For patients who do not have enough seizures for training in the first 50 days, the cutoff point was assigned at the 10^th^ seizure. This was to allow cross validation to work with at least 5 folds available, and could be tuned to allow fewer seizures to be included. To construct an online pseudo-prospective forecasting method, every time a new seizure appeared, the data leading up to that seizure was added to the training dataset and the model was retrained in order to update the model with new data.

After considering and implementing a variety of other classifiers, XGBoost was selected as the classifier in this project ^41^. XGBoost is a tree-based algorithm which is developed based on boosting ^42^. It is proven to be robust in an imbalanced scenario which is the nature of the seizure forecasting problem, as the ictal stage is extremely rare compared to the inter-ictal stage. In addition, the boosting algorithm provides automatic feature selection, which helps to select useful features and channels. A potential problem is that it has many hyper-parameters to tune, but this can be solved by combining model selection and cross validation.

Although XGBoost is able to select features automatically by sub-setting features, it would be beneficial to select features prior to feeding into the model. Due to the fact that we have a limited number of positive cases, compared to the number of features we have. Moreover, as some of the channels were not able to generate features that can provide value in prediction, it would be better to remove these features as well. As a result, every time after a new seizure, a XGBoost model with full features was trained first, and top features were then selected using the feature importance derived from the full feature model. New models would then be trained with reduced features.

The cross-validation approach was modified to guarantee each split contains both negative and positive case while keeping the temporal order. Methodology of the cross-validation approach is detailed in the supplementary material.

Since the evaluation continuously trained and validated the model as new seizures were encountered, it was crucial to keep the process efficient and keep the computational cost low. Thus, Bayesian Optimisation was used to improve the efficiency when searching for the optimal hyperparameters ^43–45^. The Bayesian Optimisation was initialised five times and kept searching for 10 times to determine the best combination. The area under the ROC curve (ROC_AUC; the ROC curve plots sensitivity versus specificity) was used for scoring, and it was based on the customised validation method we have proposed. In addition to reporting the AUC, clinically relevant measures of sensitivity (proportion of seizures correctly predicted) and proportion of time in low risk (time in low risk state divided by recording duration) were also reported for optimal thresholds obtained on the training data.

Six cases were considered when building the models with the use of different features: 1. Only using NMM features estimated by the LSTM filter. 2. Only using Univariate features. 3. Only using Spike Rate Feature. 4. Only using HFA features. 5. ACFW, Variance, and Spike Rate features combined. 6. ACFW, Variance, NMM, and Spike Rate features combined.

## Results

To investigate the seizure forecasting capacity of different features and illustrate examples of seizure tuning of feature phases on the slow and fast cycle time-scales, seizure phase histograms of the autocorrelation, variance, spike rates and NMM parameter features are shown in Figure 2 for a single patient. It can be observed that most of the phases for the entire non-seizure signals are typically broadly distributed over different phase angles. However, phases for the sample prior to the seizure times are distributed differently, and in several cases are also concentrated over a small range of angles. This can be seen from the colored bars which are concentrated differently from the uncolored ones. If seizures are more tuned to a particular phase angle this indicates the associated feature will be suitable for seizure forecasting. Moreover, the less overlap there is between the seizure (colored) histogram and the non-seizure (uncolored) histogram, the more useful the feature will be for seizure forecasting. Of the features considered in Figure 2, autocorrelation on the fast time-scale shows the tightest tuning of seizures to a particular phase, while spike rates on the slow time-scale shows the worst tuning of seizures.

**Figure 1.**
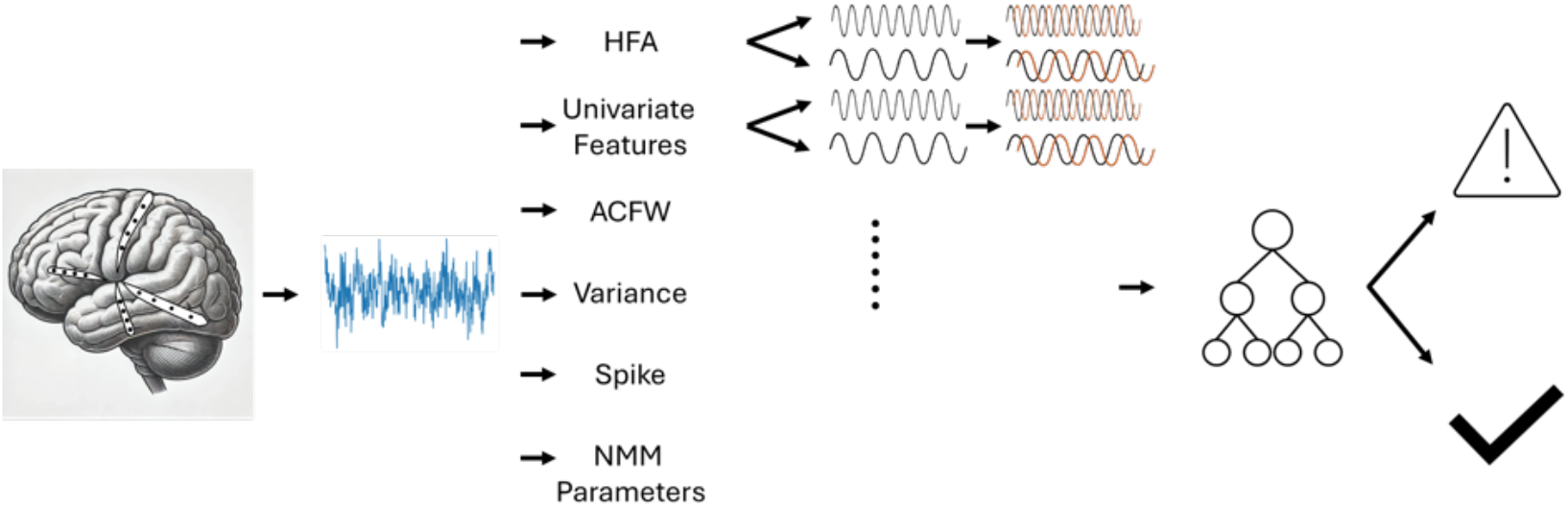
Flow chart of the paper. An implanted device collects the iEEG signal while features including High Frequency Activity (HFA), Univariate Features, Autocorrelation Function Width (ACFW), Variance, Spike Rate, and Neural Mass Model (NMM) Parameters were extracted from each iEEG channel and calculated every 2 mins. Each feature was then processed on short/fast (40 mins to 2 days) and long/slow (2 days) wideband cycles, Hilbert transformed and passed to a classifier. The classifier forecasts if there is an epileptic seizure expected in the next 2-4 minutes.

**Figure 2.**
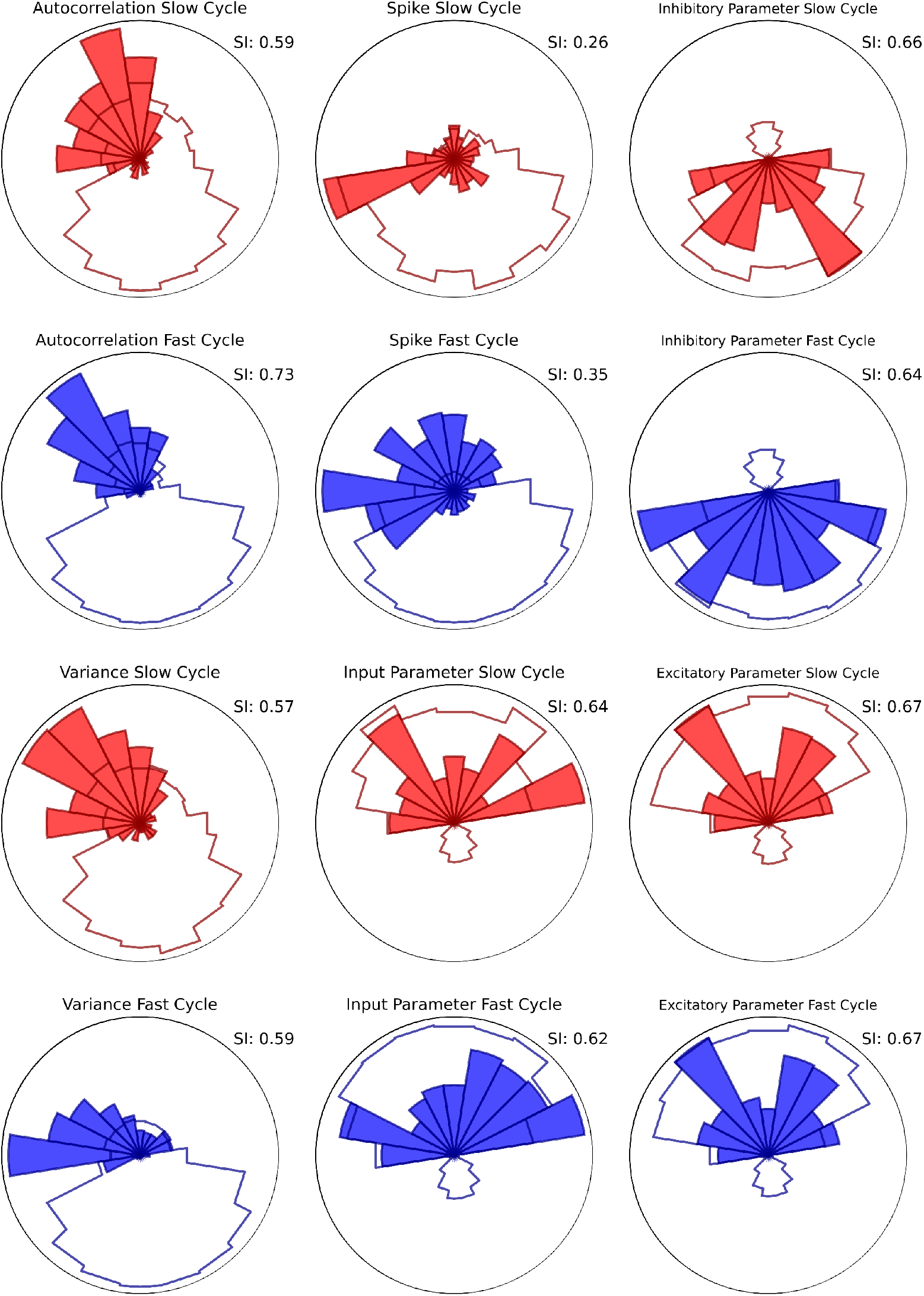
Single patient example from patient 3 autocorrelation, variance, NMM input, inhibitory parameter, and excitatory parameter normalized phase histograms for slow (red) and fast (blue) cycles. In each polar plot, the colored lines represent the normalized distribution of phases of the entire signal and the filled histogram represents the normalized distribution of phases at the sample prior to the seizure times. Phase equal to zero corresponds to the top of the black circle.

Figure shows the synchronization indices derived from the seizure phase data across the 14 different features and slow and fast time-scales for each of the subjects. As the synchronization index was calculated based on all the available data in a given channel, it shows how the feature could perform if we have access to all the data. It can be seen that the highest SI score for 12 out of 14 patients is higher than 0.8. No feature stood out as the best feature for all patients. Rather different patients had different synchronization index profiles across the features suggesting seizures were more optimally tuned to the phases of different features depending on the patient. This indicates that a machine learning approach that can combine the most appropriate features into a single seizure forecasting method is the best approach.

Figure 4 illustrates an example of the seizure forecasting method for a single patient. The raw, fast cycle, and slow cycle time series for ACFW (middle plot) and the external input parameter estimated by the LSTM filter (bottom plot) show that the signals typically peak near the seizure times (labelled by red triangles), indicating these features have a rising trend near seizures. The output result of the XGBoost model with the critical slowing down (ACFW, Variance) and NMM features is shown in the bottom of Figure 4. The probability of seizure increases when it is close to a seizure. The threshold is adjusted after adaptive retraining after each seizure, which keeps the time in low risk and the false positive rate under control.

**Figure 3.**
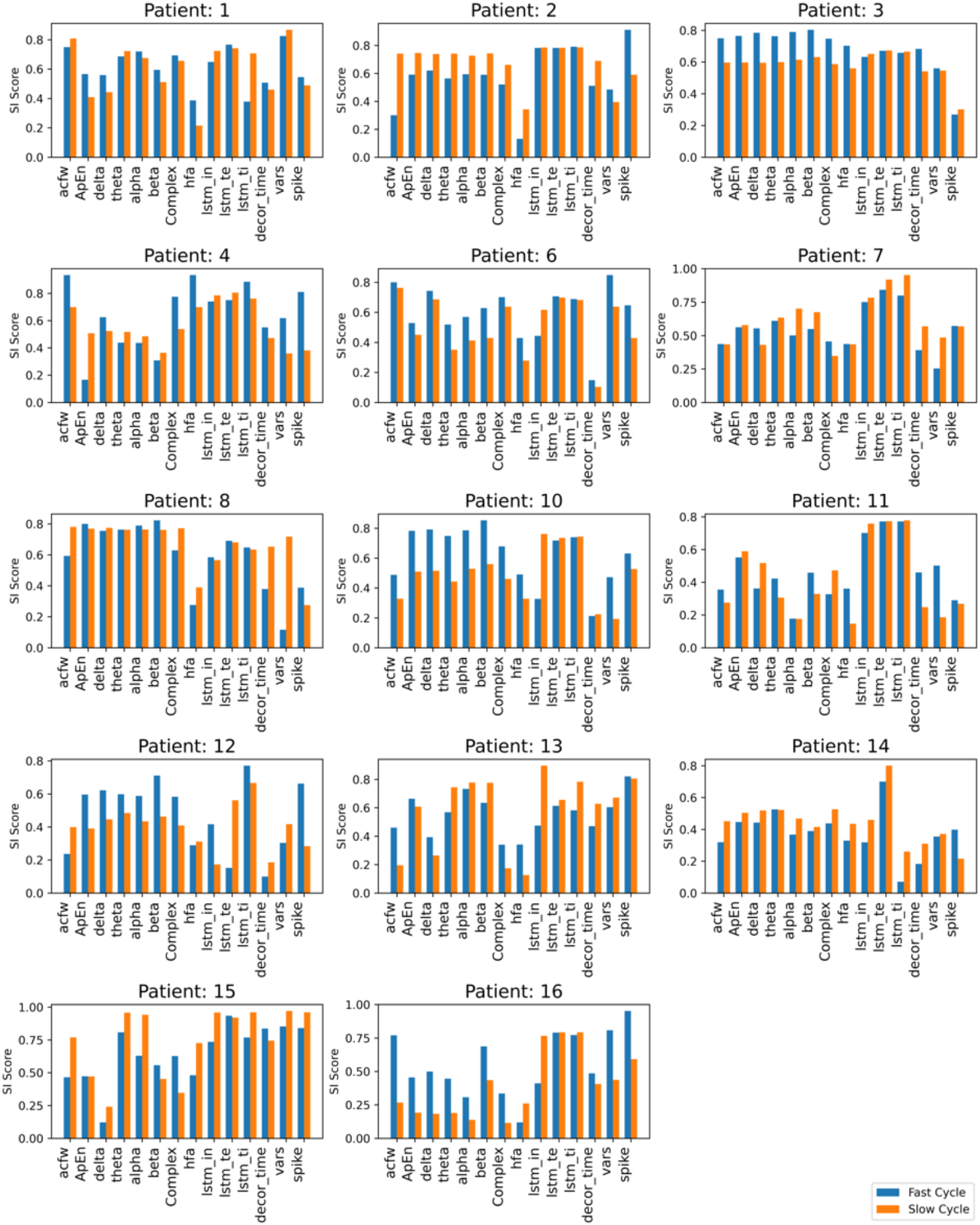
Synchronisation Index scores for each patient. All features were calculated on each patient and each channel. Only the channel with the highest SI score was shown.

**Figure 4.**
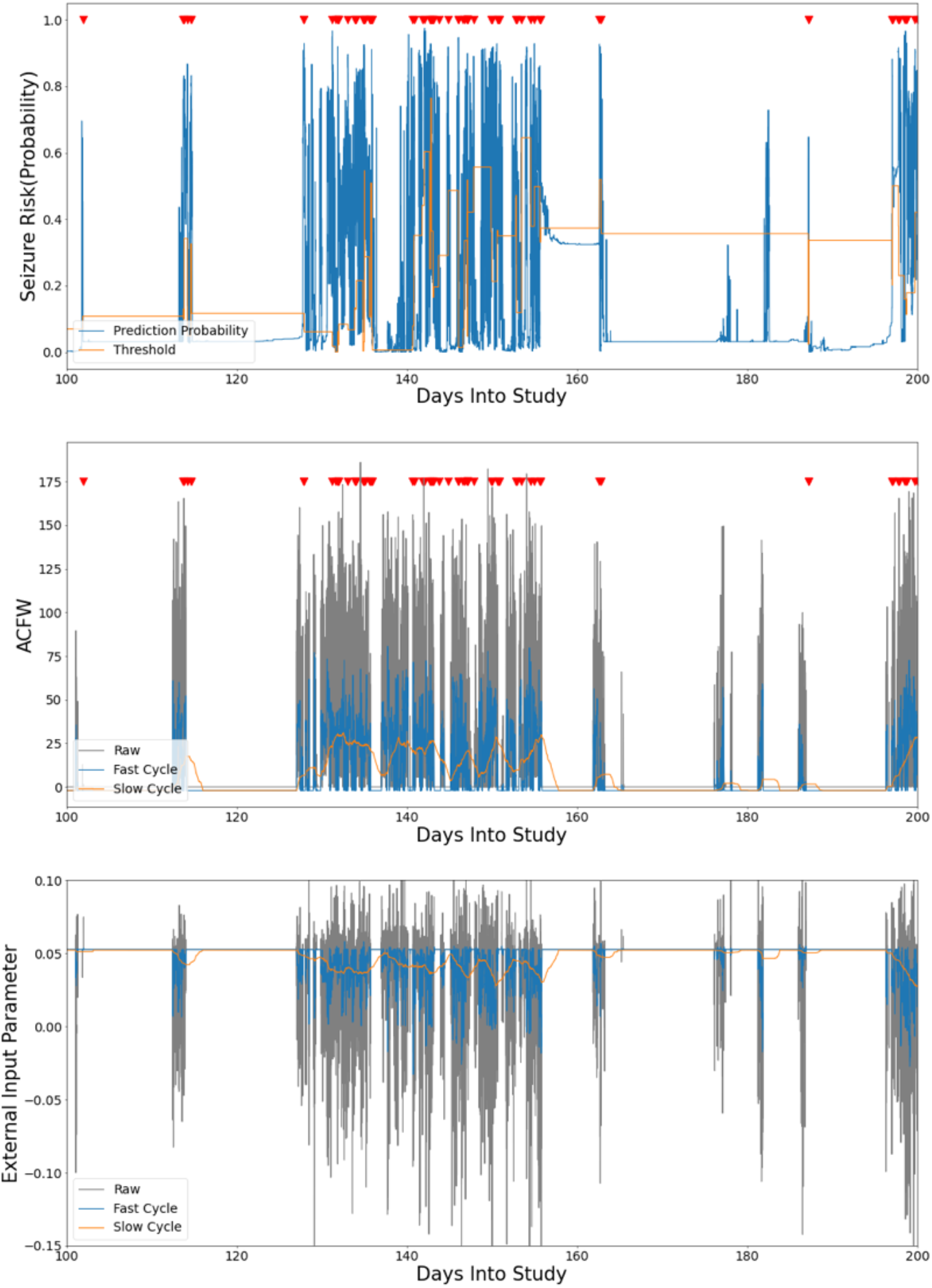
Example Seizure Risk (top), ACFW (middle), and Model External Input Parameter Estimation (bottom) time series from patient 3 over 100 days. Seizure times are labelled with red triangles. The Seizure Risk is the output of the XGBoost model, which results in a probability. The Threshold was recalculated after each new seizure. A probability (blue line) higher than the threshold (orange line) would be classified as a predicted seizure / warning, while any time lower than the threshold is not at warning stage. ACFW and External Input Parameter are the features used for the XGBoost model, where the Raw (Grey) is the raw feature value, and Fast Cycle (Blue) and Slow Cycle (Orange) are the wideband feature signals corresponding to periods of 40 mins to 2 days and 2+ days, respectively.

Testing results of time spent in low risk, sensitivity and AUC across all patients are captured in Table 1. Some approaches with one feature only can also achieve a good overall result in terms of AUC for some patients, but it is usually not easy for them to perform well for patients who have a large number of seizures. The other combined approach without using NMM parameters can achieve an average AUC of 0.8, which is close to the best approach. The best approach ACFW+Vars+NMM+Spike has a better overall result with an average AUC of 0.83.

**Table 1.** Results of the six cases. Low Risk refers to the proportion of time spent below the decision threshold for each patient, while sensitivity refers to the proportion of seizures correctly predicted. The best model (ACFW+Vars+NMM+Spike Rate) has been statistically proven to be better than chance with p-value smaller than 0.001 using the single sample Hanley & McNeil method ^46^.

| Patient | # of Seizures | NMM Only |  |  | Univariate Only |  |  | Spike Rate Only |  |  |
| --- | --- | --- | --- | --- | --- | --- | --- | --- | --- | --- |
|  |  | Low Risk | Sensitivity | AUC | Low Risk | Sensitivity | AUC | Low Risk | Sensitivity | AUC |
| 1 | 148 | 0.88 | 0.83 | 0.86 | 0.81 | 0.90 | 0.85 | 0.76 | 0.83 | 0.86 |
| 2 | 32 | 0.71 | 0.59 | 0.65 | 0.83 | 0.59 | 0.71 | 0.87 | 0.59 | 0.65 |
| 2 | 372 | 0.76 | 0.88 | 0.82 | 0.77 | 0.90 | **0.84** | 0.69 | 0.88 | 0.82 |
| 4 | 22 | 0.90 | 0.50 | 0.70 | 0.80 | 0.75 | 0.77 | 0.88 | 0.50 | 0.70 |
| 6 | 100 | 0.79 | 0.78 | 0.78 | 0.78 | 0.79 | 0.79 | 0.91 | 0.78 | 0.78 |
| 7 | 308 | 0.70 | 0.88 | 0.79 | 0.78 | 0.79 | 0.79 | 0.82 | 0.88 | 0.79 |
| 8 | 466 | 0.64 | 0.76 | 0.70 | 0.67 | 0.80 | **0.74** | 0.68 | 0.76 | 0.70 |
| 10 | 204 | 0.72 | 0.79 | 0.75 | 0.71 | 0.80 | 0.76 | 0.79 | 0.79 | 0.75 |
| 11 | 540 | 0.67 | 0.83 | 0.75 | 0.63 | 0.85 | 0.74 | 0.57 | 0.88 | 0.72 |
| 12 | 464 | 0.76 | 0.77 | 0.77 | 0.75 | 0.75 | 0.75 | 0.81 | 0.84 | 0.82 |
| 13 | 13 | 0.82 | 0.67 | 0.74 | 0.95 | 0.00 | 0.48 | 0.97 | 0.67 | **0.82** |
| 14 | 498 | 0.63 | 0.74 | 0.68 | 0.65 | 0.71 | 0.68 | 0.68 | 0.75 | 0.72 |
| 15 | 12 | 0.98 | 0.50 | 0.74 | 0.99 | 0.50 | 0.75 | 0.97 | 0.00 | 0.49 |
| 16 | 77 | 0.77 | 0.72 | 0.74 | 0.84 | 0.78 | 0.81 | 0.91 | 0.93 | **0.92** |
| Average | 233 | 0.77 | 0.73 | 0.75 | 0.78 | 0.71 | 0.75 | 0.81 | 0.72 | 0.75 |
| Median | 176 | 0.76 | 0.77 | 0.75 | 0.78 | 0.79 | 0.75 | 0.82 | 0.79 | 0.77 |
| Patient | # of Seizures | HFA Only |  |  | ACFW+Vars+Spike Rate |  |  | ACFW+Vars+NMM+Spike R. |  |  |
|  |  | Low Risk | Sensitivity | AUC | Low Risk | Sensitivity | AUC | Low Risk | Sensitivity | AUC |
| 1 | 148 | 0.66 | 0.67 | 0.66 | 0.88 | 0.79 | 0.83 | 0.90 | 0.86 | **0.88** |
| 2 | 32 | 0.50 | 0.50 | 0.50 | 0.88 | 0.82 | **0.85** | 0.90 | 0.77 | 0.84 |
| 2 | 372 | 0.72 | 0.84 | 0.78 | 0.73 | 0.91 | 0.82 | 0.75 | 0.90 | 0.83 |
| 4 | 22 | 0.43 | 0.58 | 0.50 | 0.90 | 0.83 | **0.87** | 0.85 | 0.85 | 0.85 |
| 6 | 100 | 0.71 | 0.79 | 0.73 | 0.91 | 0.80 | **0.86** | 0.85 | 0.85 | 0.85 |
| 7 | 308 | 0.52 | 0.89 | 0.71 | 0.78 | 0.80 | 0.79 | 0.78 | 0.82 | **0.80** |
| 8 | 466 | 0.64 | 0.77 | 0.71 | 0.64 | 0.81 | 0.73 | 0.67 | 0.79 | 0.73 |
| 10 | 204 | 0.54 | 0.73 | 0.63 | 0.82 | 0.88 | 0.85 | 0.82 | 0.89 | **0.85** |
| 11 | 540 | 0.63 | 0.83 | 0.73 | 0.66 | 0.86 | 0.76 | 0.69 | 0.85 | **0.77** |
| 12 | 464 | 0.70 | 0.68 | 0.69 | 0.83 | 0.85 | **0.84** | 0.82 | 0.83 | 0.82 |
| 13 | 13 | 0.67 | 0.00 | 0.34 | 0.97 | 0.33 | 0.65 | 0.94 | 0.67 | 0.80 |
| 14 | 498 | 0.57 | 0.68 | 0.62 | 0.70 | 0.76 | 0.73 | 0.73 | 0.75 | **0.74** |
| 15 | 12 | 0.85 | 1.00 | 0.92 | 0.91 | 0.50 | 0.71 | 0.97 | 1.00 | **0.99** |
| 16 | 77 | 0.70 | 0.64 | 0.67 | 0.90 | 0.84 | 0.87 | 0.89 | 0.79 | 0.84 |
| Average | 233 | 0.63 | 0.69 | 0.66 | 0.82 | 0.77 | 0.80 | 0.83 | 0.83 | **0.83** |
| Median | 176 | 0.65 | 0.71 | 0.68 | 0.86 | 0.82 | 0.83 | 0.84 | 0.84 | **0.83** |

## Discussion

Overall, the results indicate that a method that can selectively and adaptively combine all of the considered features, in particular ‘ACFW+Vars+NMM+Spike Rate’, works best for most patients and performs no worse than the single feature models for all patients. Considering the more mechanistically interpretable features, it was found that the proposed method can achieve a good performance of around 0.75 AUC using NMM features alone. Even though the performance is better with ACFW and Variance, these two features are less biophysically interpretable, while NMM features can be explored further with respect to their dynamical properties in different regions of the parameter space.

With the results of all cases, it was observed that it is possible to find a method for a patient-specific seizure warning system without having to find the best channel. As the best channel must be chosen prior to applying the methodology, determining the channel to be used is usually difficult. The proposed method does not require the channel selection process to be included prior to the implementation of the model. This can be further studied patient-specifically to find the brain area that is suitable for prediction. Features especially the NMM features can be used to derive insights in the neurophysiologically interpretable parameter space.

Although the results were not shown here, compared to feeding the features calculated on the fast and slow time scales straight into the XGBoost classifier, feeding the feature phases after the Hilbert Transform provided better performance. This could imply seizure forecasting benefits from time dependency, as the Hilbert Transform presents the signal phase at a certain time step which makes use of the signal at other time points. It might be better if further work can try time dependent classifier or forecasting models that take the timesteps before the current one into consideration during training and prediction, since XGBoost makes prediction only at one timestep without using any other timestep. In this way, the Hilbert Transform might not be necessary, and the raw features on different time-scales can be directly used. However, as the nature of the data is highly imbalanced, it is difficult to implement such methods like Transformers, as they typically require a large amount of data. Although the data were recorded for up to three years, the number of positive cases (pre-ictal stage) is limited.

Every feature extracted from the raw signal can show the fast and slow cycles after Hilbert Transform. However, different features’ rhythms are more strongly or weakly associated with seizures for different patients. Generally fast cycles showed stronger SI values, but that was not always the case. Nevertheless, combining stronger or weaker rhythms into a forecasting model can still generate robust performance. Given a seizure is always going to be a rare event, and the seizure prediction task will always have a low number of examples for a single patient, it is necessary to only include features that can show good performance for the patient. The best models have included features that generally showed a better performance (ACFW, Vars, Spike Rate, and NMM features) with slow and fast cycles. Future works can focus on the instantaneous features as well as bi-or multi-variate features that combine different channels ^15^. On the other hand, features with cycles might also indicate the mechanism of seizures for the specific patient.

The results using the HFA feature were different from an original study using the same data ^47^. This is due to the sampling being used here being based on the first 50 days of the data, instead of the whole period for each patient. In addition, sampling was done by randomly selecting 300 segments to reduce the computational cost, which is also different. Considering the changes made to the original method, the average AUC was 0.66, which is lower than the AUC reported in the original experiment.

Compared to the original critical slowing down seizure forecasting study^13^ which reported an average sensitivity of 0.77 with 0.87 proportion of time in low risk using the combination of ‘ACFW+Vars+Spike Rate’, the best model (‘ACFW+Vars+NMM+Spike Rate’) achieved a similar result. On top of this, it eliminated the need of selecting the best channel and features prior to model building. Moreover, for the same data a recent study using HFA reported an average AUC around 0.73 ^47^, while another study using LSTM has reported an average AUC of 0.75 ^48^. Given the best model here achieved an average AUC of 0.83, this suggests it represents the current state of the art for this dataset and future work on long-term recordings from other patients will be needed to verify how well this approach generalises to other patients.

## Data Availability

Seizures and some segments of the data used in this study are currently publicly available on the online platform Epilepsyecosystem. Other segments of the data can be made available upon reasonable request. The data that support the findings of this study are openly available at the following URL/DOI: https:// doi.org/10.26188/5b6a999fa2316.

## Data availability

Seizures and some segments of the data used in this study are currently publicly available on the online platform Epilepsyecosystem^12^. Other segments of the data can be made available upon reasonable request. The data that support the findings of this study are openly available at the following URL/DOI: https://doi.org/10.26188/5b6a999fa2316.

## Funding

Research supported by Australian Research Council (DP200102600).

## Competing interests

The authors report no competing interests.

## Notes

### Competing Interest Statement

The authors have declared no competing interest.

### Author Declarations

Human Research Ethics Committee, St. Vincent's Hospital, Melbourne gave ethical approval for this work

