## Supplementary material for "Seizure Forecasting with Multiple Time Scales and Features"

Author affiliations:

Full address Dept. of Data Science and AI, Faculty of Information Technology, Monash University, Clayton, VIC, Australia

### NMM

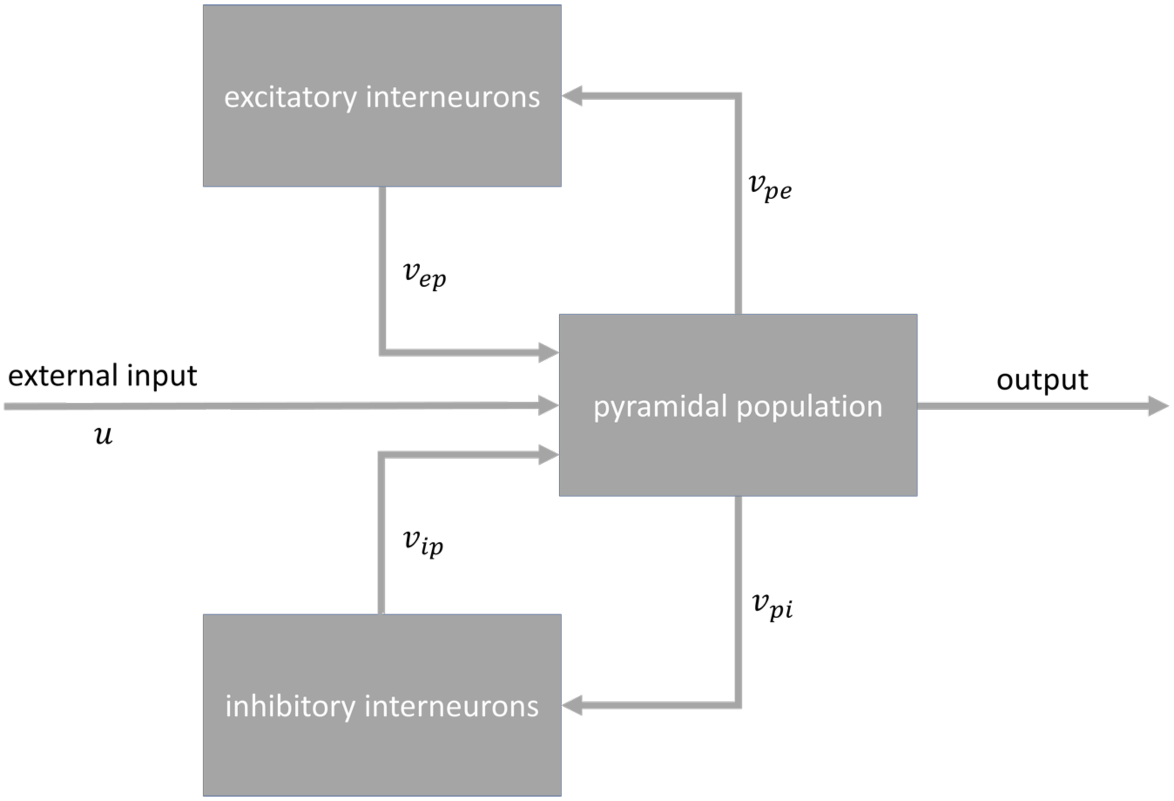


*Figure 1Structure of the Jansen & Rit neural mass model [1]. The model consists of three populations: pyramidal neurons, inhibitory interneurons and excitatory interneurons. The output of each population is a firing rate, which is transformed to changes in the mean membrane potentials of connected neural populations*

The post-synaptic potential of population m arising as a result of input from pre-synaptic population n is expressed as

$$v_{mn}(t) = \alpha_{mn} \int_{-\infty}^{t} h_{mn}(t-t')\phi\big(v_n(t')\big)dt' \quad (1)$$

where $\alpha_{mn}$ is the population averaged synaptic connectivity strength [2,3].

The convolution in equation (1) can also be written as two coupled, first order, ordinary differential equations.

$$\frac{dv_{mn}}{dt} = z_{mn} \quad (2)$$

$$\frac{dz_{mn}}{dt} = \frac{\alpha_{mn}}{\tau_{mn}}\phi_{mn} - \frac{2}{\tau_{mn}}z_{mn} - \frac{1}{\tau_{mn}^2}v_{mn} \quad (3)$$

where $\tau_{mn}$ is a lumped time constant, and m and n indicate the pre-synaptic and post-synaptic neural population, respectively. $\alpha_{mn}$ is the parameter we are mainly estimating to represent the connectivity strength between neural populations.

Furthermore, it is helpful to express the NMM in matrix-vector notation so the operations can be expressed more compactly.

$$\dot{x}(t) = Ax(t) + B\vec{\phi}\big(Cx(t)\big) \tag{4}$$

$$y(t) = Hx(t) + v(t) \tag{5}$$

where H is the observation matrix, $v(t) \sim N(0, R)$ is the observation noise and $y(t)$ is the membrane potential of the pyramidal population, which is considered to be the contributor to the generation of the EEG signal in our model.

In order to show the set of parameters for estimation, we define a vector of parameters as $\theta = \left[u\ \alpha_{pe}\ \alpha_{pi}\ \alpha_{ip}\ \alpha_{ep}\right]^T$. This set corresponds to the input $\mu$ to the model and the population averaged connectivity strength parameters. Moreover, we assume the time constants of the model to be constant to simplify the estimation problem. The parameter vector is combined with the state vector X to from the augmented state vector,

$$\xi = [\mathrm{X}^{\mathrm{T}}\theta^{\mathrm{T}}]^{T} \tag{6}$$

The augmented state-space model is then

$$\xi_t = A_\theta \xi_{t-1} + B_\theta \phi(C_\theta \xi_{t-1}) + W_{t-1} \tag{7}$$

where $W_t$ is Gaussian noise.

#### LSTM Filter

A structure with 128 and 32 neurons in layers 1 and 2, respectively was proposed to construct the LSTM model [3]. To link an LSTM neural network with the NMM, we customised the loss function to allow the LSTM model, so that the LSTM model can learn to follow the mathematical relationship of the NMM. Since the LSTM is able to produce the values of both the observation and the state for the current timestep t given the state of the last timestep $t - 1$, it is possible to compare the state at timestep t with the state provided by the NMM, so we can know the difference between these two predictions. If we link the NMM to the loss function, the LSTM model would learn to minimise the difference.

The basic loss function for a regression problem at a single timestep is defined as

$$\text{Squared Error} = \left[\left(\xi_t - \hat{\xi}_t\right)^2,\ \left(y_t - H\hat{\xi}_t\right)^2\right] \tag{8}$$

where $\xi$ is the augmented state, y is the observation of the training data and $\hat{\xi}$ and $H\hat{\xi}_t$ are the augmented state and observation predicted by the LSTM model, respectively.

The model error is then added, which is the error between the state of the LSTM model and the state generated by the NMM:

$$\text{Model Error} = \left[\left(\hat{\xi}_t - \left(A_\theta \hat{\xi}_{t-1} + B_\theta \phi\left(C_\theta \hat{\xi}_{t-1}\right)\right)\right)^2, 0\right] \tag{9}$$

By minimising the model error, the LSTM model will learn to follow the mathematical expression of the NMM, since the training has to minimise the error between its own prediction and the state generated by the NMM.

Lastly, as we consider the connectivity strength parameters to be slowly changing parameters compared to the membrane potential and simulated EEG signals, we have to control the rate of change for these parameters. We can add the standard deviation of the parameters to the loss function to limit the changing rate. However, the parameters also have to be adjusted rapidly when they are too far away from the NMM. Thus, the standard deviation is combined with the model error as

$$\text{Std} = s(\alpha)\left[\left(\hat{\xi}_t - \left(A_\theta \hat{\xi}_{t-1} + B_\theta \phi\left(C_\theta \hat{\xi}_{t-1}\right)\right)\right)^2, 0\right] * k \tag{10}$$

$$s(\alpha) = [0, \dots 0, std(\alpha), 0]^T \tag{11}$$

where $\alpha$ is the vector of the four connectivity strengths parameters and k is an adjustable weight that is set to 0.1 as the default value.

The final loss function is then the summation of equation (8), (9), and (10).

#### Cross Validation

The traditional stratified k-fold cross validation approach is not suitable, as the data has a temporal order. Moreover, traditional forward chaining is also not a valid approach in our scenario, as seizures are extremely rare, it cannot guarantee there is at least one positive case in each split. Thus, we have customized a cross validation approach, which splits the data based on the positive cases. The time of labels were identified first. If there were two positive labels contiguous with each other, the one that came first chronologically would be kept, and the one that came later would be ignored in the splitting process. This is to guarantee each split contains negative cases. Folds would then be split based on the time of positive cases, which means each fold would have different length, but would keep similar number of positive cases. In order to follow the nature of the time series data, the concept of forward chaining [4] was employed, so in the $n^{th}$ split, the first n split would be used as training data, while the $(n+1)^{th}$ split would be used for validation.

#### Threshold Optimisation

It is also important to note that the positive case is extremely rare compared to the normal state, and in some cases, the normal stage data would show very similar pattern to preictal stage data, but does not lead to a seizure. In other words, it is important to update the model to allow and control false positives. To solve for this issue, the threshold for determining prediction results was adjusted every time a new model was established. The model was fitted on the splits except for the last one, while the last split was used only for adjusting the threshold. To adjust the threshold, the ROC curve was first calculated using the prediction and the labels for the last split, then calculated the G-mean ($g$) for each threshold [5]. The threshold with the largest G-mean was then found to be the optimal threshold.

$$g = \frac{a^+}{a^-} \tag{12}$$

where $a^+$is the true positive rate, and $a^-$is the false positive rate.
